# Challenges, innovations and considerations for the use of tongue swabs in *Mycobacterium tuberculosis* complex detection

**DOI:** 10.1101/2025.05.18.25327877

**Authors:** Anura David, Keneilwe Peloakgosi-Shikwambani, Zanele Nsingwane, Violet Molepo, Wendy Stevens, Lesley Scott

## Abstract

**Background:** Sputum-based testing has long been the cornerstone of tuberculosis (TB) diagnosis, but emerging specimen types like tongue swabs (TS) offer additional approaches for detecting *Mycobacterium tuberculosis* complex (MTBC). This two-phase study evaluated the impact of storage and transport conditions, along with laboratory processing modifications, to improve MTBC detection using TS tested on quantitative PCR (qPCR) and Xpert MTB/RIF Ultra (Ultra).

**Methods:** Participants with positive Ultra sputum were enrolled at a healthcare facility in Johannesburg, South Africa. In Phase one, five serial TS were collected per participant and transported “dry” at 2-8°C. In Phase two, seven TS were collected and stored “dry,” in Tris-EDTA (TE) buffer or PrimeStore® Molecular Transport Medium (MTM) and subjected to different storage temperatures (−80°C or 37°C) before laboratory testing.

**Results:** Results using qPCR and Ultra demonstrated that MTBC detection from serially collected TS was possible but sporadic, particularly in individuals with lower bacillary loads. Vortex bead beating (VBB) improved detection compared to heat lysis alone. Storage at −80°C was shown to be a viable option for longer term storage while short-term storage at 37°C was feasible. Ultra testing showed improved detection when TS were processed with diluted SR buffer.

**Conclusions:** TS offer a viable, though variable, specimen type for MTBC detection. Optimizing processing methods, such as incorporating VBB and diluted SR buffer, and selecting appropriate storage conditions can enhance diagnostic performance and support broader implementation in diverse settings.

## Introduction

The diagnosis of tuberculosis (TB) typically relies on sputum and other respiratory specimens for bacteriological confirmation (1). However, additional specimen types such as urine (2), stool (3) and more recently tongue swabs (TS) (4) have shown value in detecting *Mycobacterium tuberculosis* complex (MTBC). TS offer a non-invasive alternative for populations where sputum collection is difficult, such as children, people living with HIV (PLHIV), or individuals with paucibacillary disease.

Since 2021, our group has been exploring the utility of TS for the detection of MTBC, at a time when limited published evidence existed to inform specimen collection, processing protocols, or performance expectations. As with any novel specimen type, evaluating TS for TB diagnosis requires rigorous research and protocol optimization to understand its operational and diagnostic potential. While studies using contrived samples play an important role in early-stage development, clinical validation in real-world settings is essential to assess true diagnostic utility.

This paper presents findings from an early two-phase study that investigated whether TS performance for MTBC detection is influenced by storage and transport conditions, including temperature and buffer preservation compared to dry transport and storage. Additionally, the study assessed the impact of laboratory processing modifications on detection yield. Our results indicate that both pre-processing conditions and storage temperature affect the sensitivity of TS for MTBC detection.

## Materials and Methods

### Study design and TS collection

In this cross-sectional study, we recruited adult (≥ 18 years) participants with positive Xpert MTB/RIF Ultra (Cepheid, Sunnyvale, CA, USA) (Ultra) sputum results from the Hillbrow Community Health Centre in Johannesburg, South Africa. Participants were approached and invited to enroll in the study. Recruitment was performed over two phases. In phase 1, five serially collected TS were collected by a research nurse and in phase 2, seven serially collected swabs were collected, after obtaining written informed consent. For both phases, TS were collected when participants returned to the healthcare facility to receive their Ultra results, prior to the initiation of TB treatment. Swabs were collected in a random order rather than sequentially by swab number. TS collection followed the method described by Andama *et al.* (5) with TS collected approximately two minutes apart. Each TS was stored in a 2 mL cryovial and transported to the research laboratory in Braamfontein, Johannesburg, for testing.

#### Phase One

Recruitment took place from 19 October 2022 – 15 March 2023. Thirty-eight participants provided consent. All five TS were transported “dry” or without any buffer, at 2-8°C, to the laboratory.

#### Phase Two

Findings from Phase 1 guided the design of phase 2, during which two additional TS were collected per participant. In phase 2, we further evaluated factors such as transport and storage in Tris-EDTA [TE] [10 mM Tris-HCl containing 1 mM EDTA Na₂, pH 8.0; Merck, Johannesburg, SA]) buffer, along with an additional storage temperature (37°C), to determine (1) whether TS performance could be improved and (2) whether refrigeration is necessary for TS storage prior to laboratory processing.

From 18 May - 21 August 2023, an additional 39 participants were recruited where seven TS were serially collected from each participant. Three swabs were transported “dry,” two were transported in TE buffer and the remaining two were transported in PrimeStore® Molecular Transport Medium (MTM) (Longhorn Vaccines & Diagnostics, Bethesda, MD, USA). All TS were transported at 2-8°C to the laboratory except for those in MTM which were transported at room temperature. The two TS in MTM were transported in either 2.2 mL or 1.5 mL MTM.

### Swab processing at the laboratory

#### Phase One

Upon receipt at the laboratory, TS were processed as described in Fig 1.

**Fig 1:**
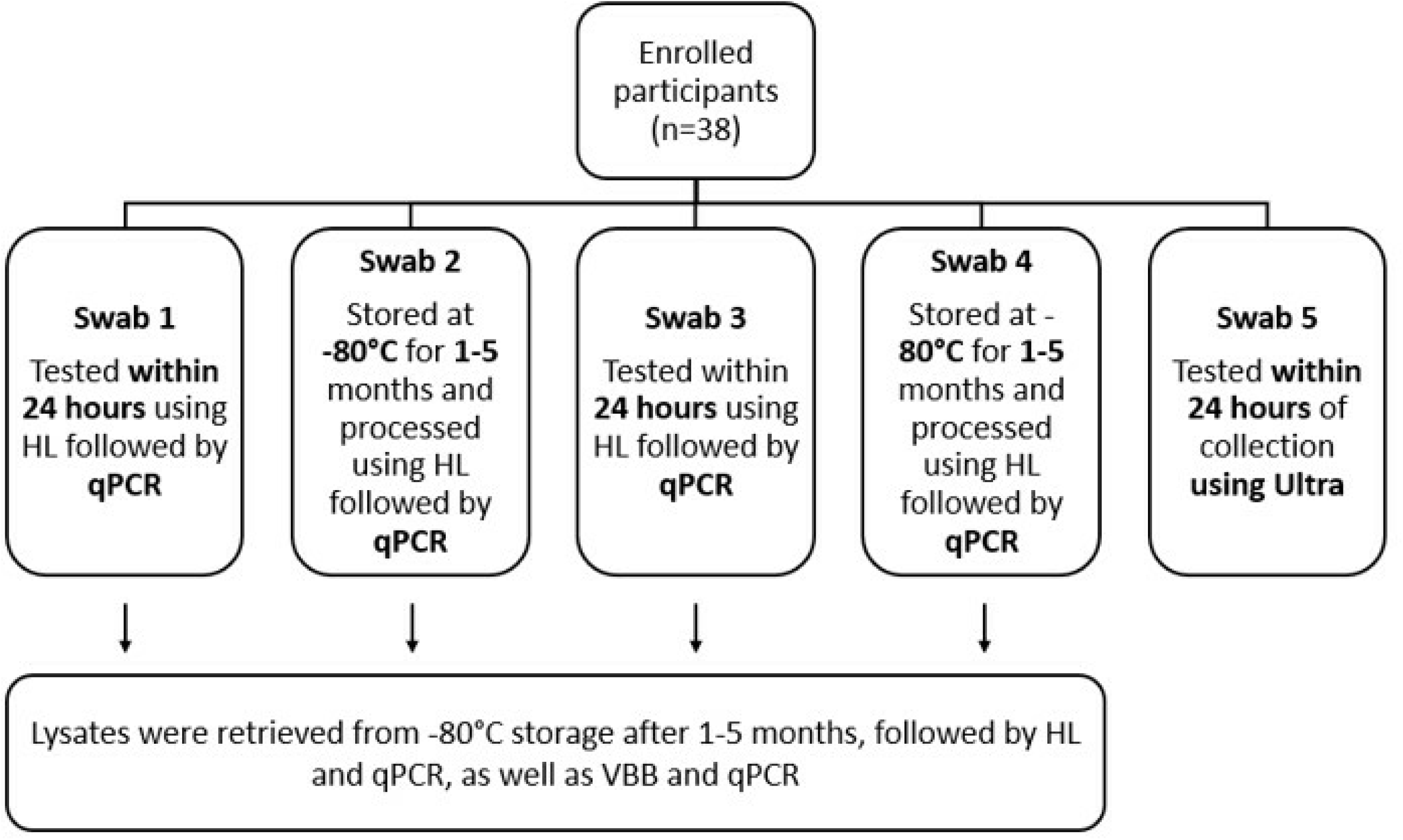
Schematic of tongue swab collection and testing strategy for participants enrolled in Phase 1 (n=38). Swabs 1, 3, and 5 were tested within 24 hours of collection using either quantitative PCR (qPCR) or Xpert MTB/RIF Ultra (Ultra). Swabs 2 and 4 were stored at −80°C for 1–5 months prior to testing by qPCR. All swabs processed by qPCR were initially subjected to heat lysis (HL); stored lysates were then subjected to an additional HL step followed by vortex bead beating (VBB).

For pre-processing of the fresh and frozen TS (Swabs 1-4), 400 µl of TE buffer was added to each cryovial containing the TS, and ysed in a heating block for 10 minutes at 95°C. The lysate was then used as a template on a real-time quantitative polymerase chain reaction (qPCR) targeting the *IS6110* and *IS1081* genes, using primer and probe design from the Quantigen group (6) with testing performed on the QuantStudio 5 system (Thermo Fisher Scientific, Waltham, MA, USA). Thresholds of 0.06 and 0.05 were used for the *IS6110* and *IS1081* genes, respectively to standardize cycle threshold (Ct) values. If only one MTBC target amplified with a Ct ≥38, the test was interpreted as inconclusive and repeated. Lysates were frozen at −80°C for 1-5 months after which they were retrieved from storage and thawed. Heat lysis (HL) was repeated, followed by qPCR. The heat lysed TS were then subjected to 5-minute vortex bead beating (VBB) using a vortex and bead beating attachment (SI-H524, Horizontal Microtube Holder (24 tubes), Scientific Industries Inc, NY, USA). Bead beaten lysates were also tested on qPCR.

For TS Ultra testing, 2.2 mL Sample Reagent (SR) (Cepheid, Sunnyvale, CA, USA) was added to the TS, incubated at room temperature for 15 minutes and 2mL of the specimen was added to the Ultra cartridge and tested as per manufacturer instructions.

#### Phase Two

Laboratory results from phase one which showed that VBB improved detection of MTBC, compared to HL alone guided, laboratory testing for phase two. One swab was tested within 24 hours of collection, by performing VBB followed by qPCR. Two TS were stored at −80°C and another two at 37°C. After a 3-day and 7-day incubation, one TS from each storage temperature was retrieved and tested using VBB and qPCR (Fig 2). As per the phase 1 protocol, 400 µl of TE buffer was added to all “dry” TS. For biosafety consideration and to assist with lysis of the Mycobacteria, all TS were subjected to a 10-minute HL before VBB.

**Fig 2:**
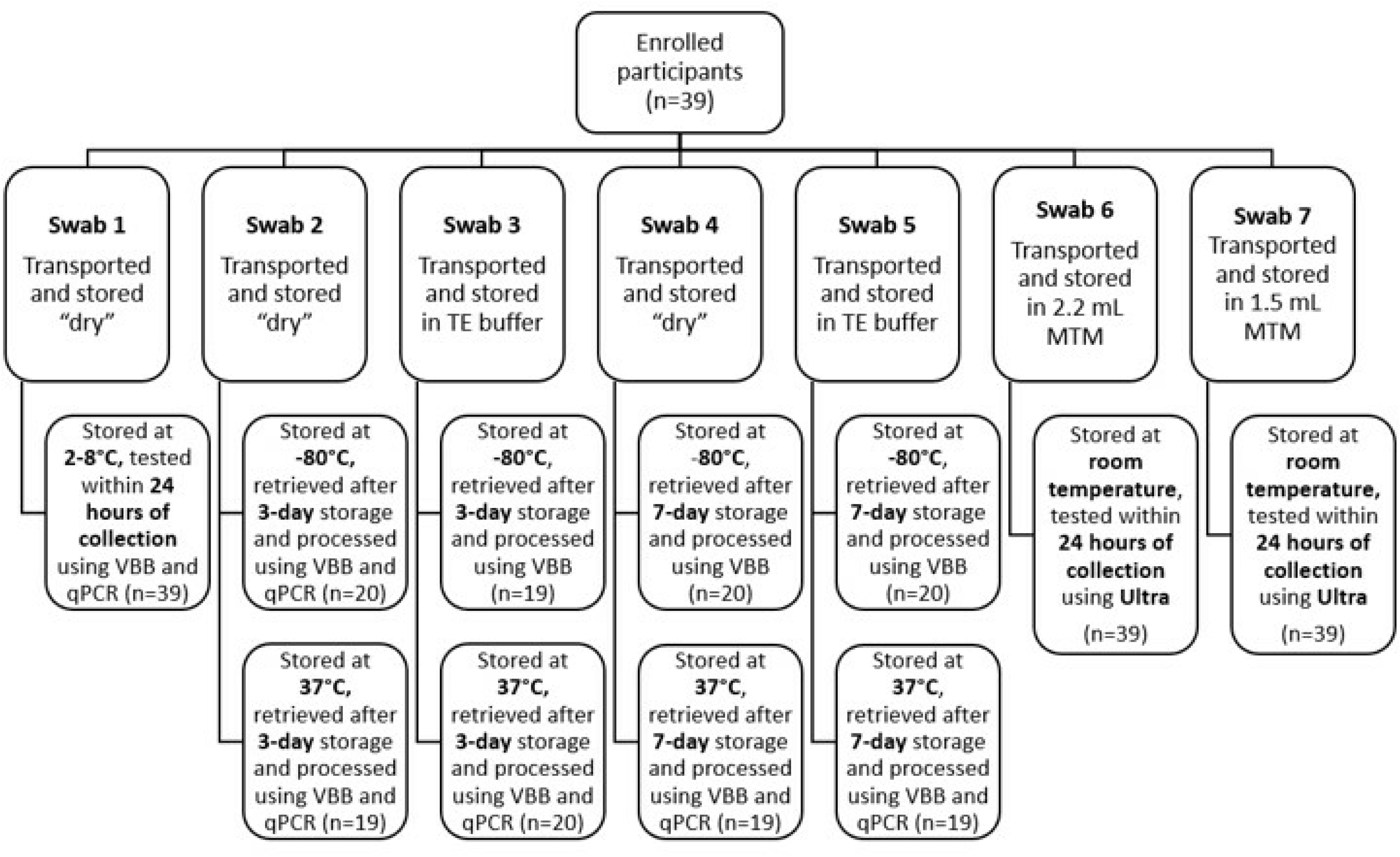
Overview of tongue swab collection, transport, storage conditions, and testing methods for participants in Phase 2 (n=39). Swabs were stored “dry,” in Tris-EDTA (TE) buffer, or PrimeStore® MTM, and tested using qPCR or Xpert Ultra after various storage durations (24 hours to 7 days) and temperatures (2–8°C, −80°C, 37°C, or room temperature).

For TS Ultra testing, 2mL MTM was removed from the vial containing 2.2 mL MTM and added directly to the Ultra cartridge. For the TS containing 1.5 mL MTM, 1.5 mL SR was added, incubated for 15 minutes at room temperature and 2mL used for Ultra testing. Once the specimen was added to the Ultra cartridge, testing was performed as per manufacturer instructions.

#### Statistical analysis

Due to small sample sizes, descriptive statistics were used to summarize participant characteristics and MTBC detection rates across specimen types, processing methods, and test platforms. Detection rates were calculated as proportions with corresponding percentages.

Comparisons of detection performance between processing methods, specifically HL versus VBB were also assessed. Differences in MTBC detection across semi-quantitative categories of the Ultra sputum assay were similarly analyzed.

Cycle threshold values obtained from qPCR testing were compared between pre- and post-storage conditions, and between HL and VBB treatment.

## Results

### Phase One

Among participants with valid results for all tests, all five TS tested positive by qPCR and Ultra in 5/38 (13%) participants (Table 1). All five participants had “high” or “medium” semi-quantitative results on their Ultra sputum assay. For other participants, MTBC detection across the five TS was sporadic. When tested using qPCR, TS tested within 24 hours of collection, and when using HL alone, MTBC was detected on 53/152 (35%) specimens, while processing with VBB detected MTBC on 36/76 (47%) specimens. Ct values remained consistent for specimens lysed using heat alone, both before and after storage (Table S1) while VBB treatment led to a 1–4 cycle reduction in Ct values (Fig 3). Repeat HL did not improve assay sensitivity since MTBC detection rates were similar. For Ultra TS testing, MTBC detection varied across the Ultra sputum semi-quantitative range with 18/38 (47%) testing positive for MTBC. A total of 5/38 (13%) Ultra TS specimens produced unsuccessful results.

**Fig 3:**
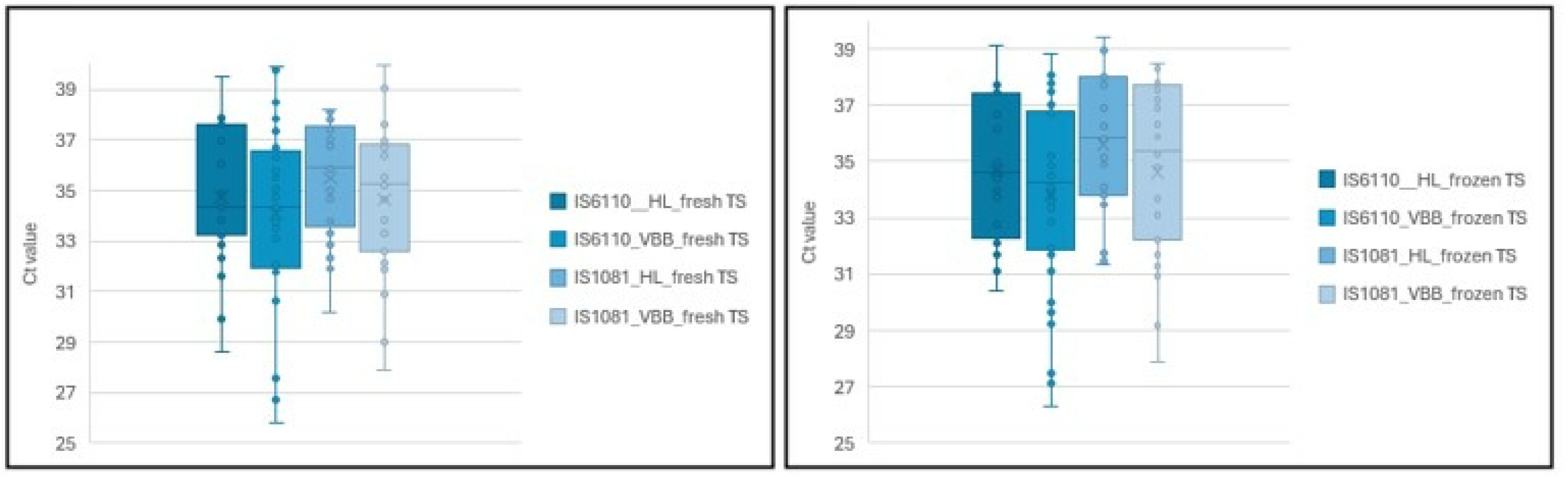
Ct values for insertion elements (*IS*6110 and *IS*1081) obtained on qPCR between tongue specimens that were heat lysed only compared with a combination of heat lysis and vortex beat beating. Figure 3A represents TS that were processed “fresh” or within 24 hours of collection and Figure 3B represents TS that were processed after freezing

**Table 1:**
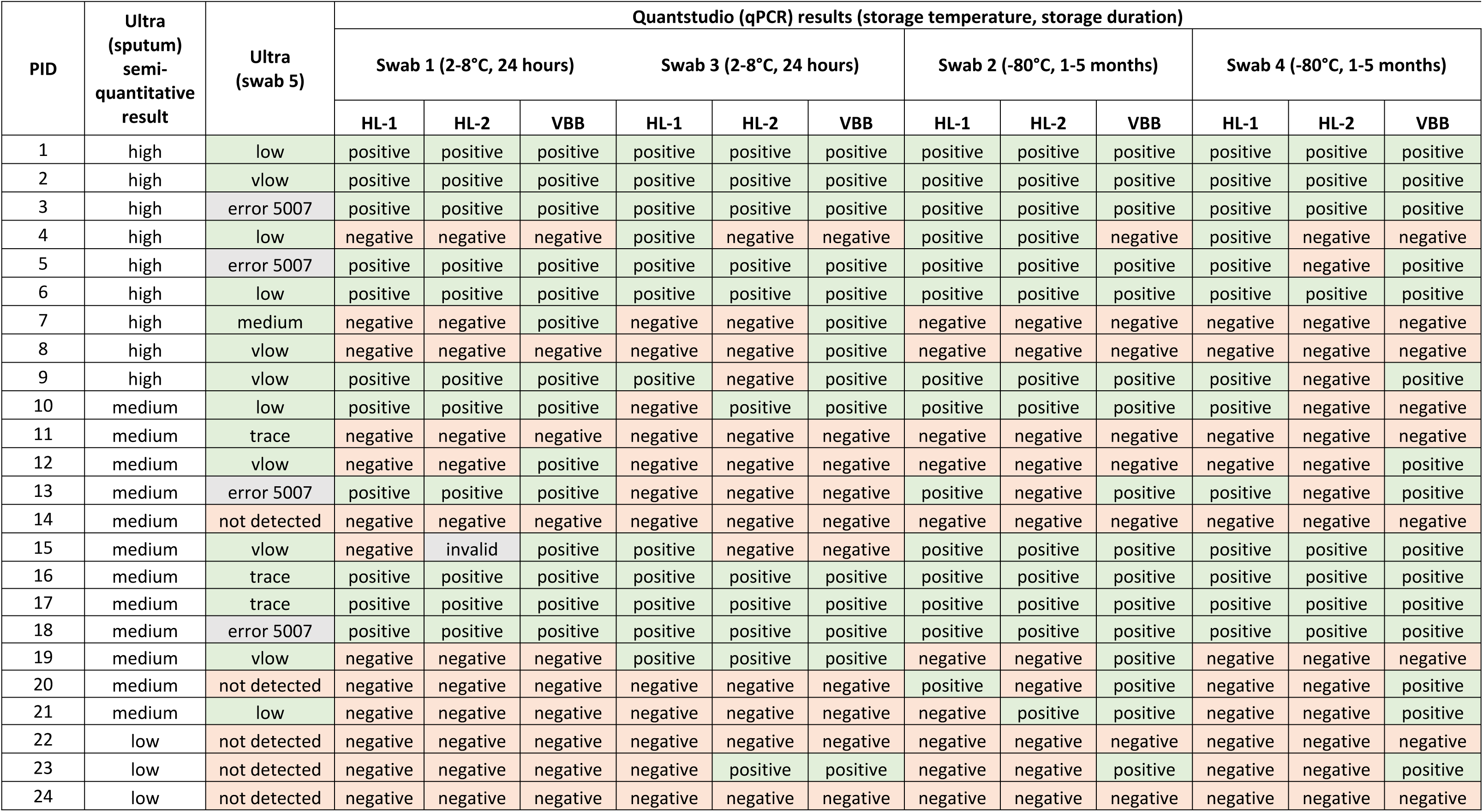
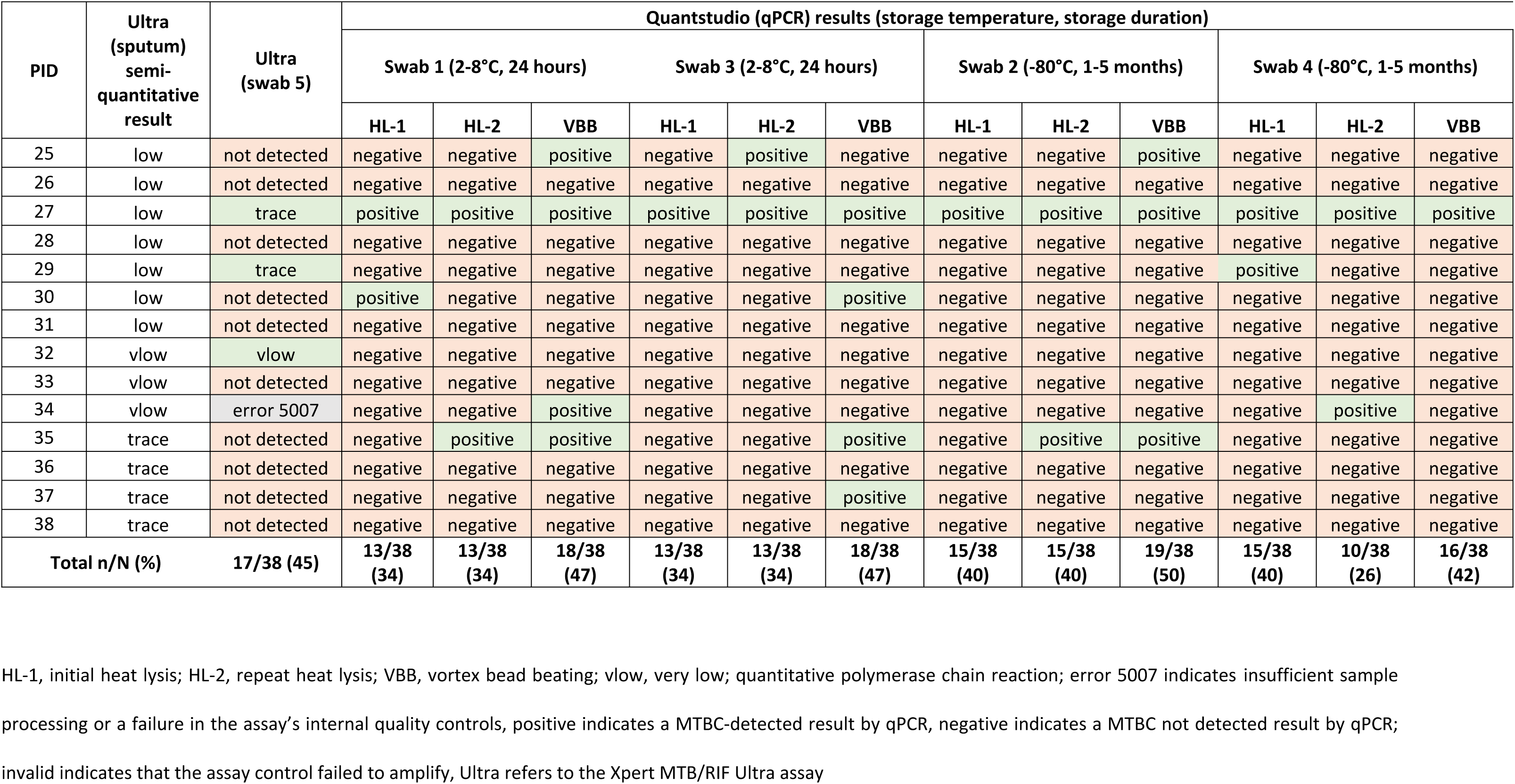
Ultra semi-quantitative and qPCR results for tongue swabs from participants recruited during phase one.

| PID | Ultra<br>(sputum)<br>semi-quantitative<br>result | Ultra<br>(swab 5) | Quantstudio (qPCR) results (storage temperature, storage duration) |  |  |  |  |  |  |  |  |  |  |  |
| --- | --- | --- | --- | --- | --- | --- | --- | --- | --- | --- | --- | --- | --- | --- |
|  |  |  | Swab 1 (2-8°C, 24 hours) |  |  | Swab 3 (2-8°C, 24 hours) |  |  | Swab 2 (-80°C, 1-5 months) |  |  | Swab 4 (-80°C, 1-5 months) |  |  |
|  |  |  | HL-1 | HL-2 | VBB | HL-1 | HL-2 | VBB | HL-1 | HL-2 | VBB | HL-1 | HL-2 | VBB |
| 1 | high | low | positive | positive | positive | positive | positive | positive | positive | positive | positive | positive | positive | positive |
| 2 | high | vlow | positive | positive | positive | positive | positive | positive | positive | positive | positive | positive | positive | positive |
| 3 | high | error 5007 | positive | positive | positive | positive | positive | positive | positive | positive | positive | positive | positive | positive |
| 4 | high | low | negative | negative | negative | positive | negative | negative | positive | positive | negative | positive | negative | negative |
| 5 | high | error 5007 | positive | positive | positive | positive | positive | positive | positive | positive | positive | positive | negative | positive |
| 6 | high | low | positive | positive | positive | positive | positive | positive | positive | positive | positive | positive | positive | positive |
| 7 | high | medium | negative | negative | positive | negative | negative | positive | negative | negative | negative | negative | negative | negative |
| 8 | high | vlow | negative | negative | negative | negative | negative | positive | negative | negative | negative | negative | negative | negative |
| 9 | high | vlow | positive | positive | positive | positive | negative | positive | positive | positive | positive | positive | negative | positive |
| 10 | medium | low | positive | positive | positive | negative | positive | positive | positive | positive | positive | positive | negative | negative |
| 11 | medium | trace | negative | negative | negative | negative | negative | negative | negative | negative | negative | negative | negative | negative |
| 12 | medium | vlow | negative | negative | positive | negative | negative | negative | negative | negative | negative | negative | negative | positive |
| 13 | medium | error 5007 | positive | positive | positive | negative | negative | negative | positive | negative | positive | positive | negative | positive |
| 14 | medium | not detected | negative | negative | negative | negative | negative | negative | negative | negative | negative | negative | negative | negative |
| 15 | medium | vlow | negative | invalid | positive | positive | negative | negative | positive | positive | positive | positive | positive | positive |
| 16 | medium | trace | positive | positive | positive | positive | positive | positive | positive | positive | positive | positive | positive | positive |
| 17 | medium | trace | positive | positive | positive | positive | positive | positive | positive | positive | positive | positive | positive | positive |
| 18 | medium | error 5007 | positive | positive | positive | positive | positive | positive | positive | positive | positive | positive | positive | positive |
| 19 | medium | vlow | negative | negative | negative | positive | positive | positive | negative | negative | positive | negative | negative | negative |
| 20 | medium | not detected | negative | negative | negative | negative | negative | negative | positive | negative | positive | negative | negative | positive |
| 21 | medium | low | negative | negative | negative | negative | negative | negative | negative | positive | positive | negative | negative | positive |
| 22 | low | not detected | negative | negative | negative | negative | negative | negative | negative | negative | negative | negative | negative | negative |
| 23 | low | not detected | negative | negative | negative | negative | positive | positive | negative | negative | positive | negative | negative | positive |
| 24 | low | not detected | negative | negative | negative | negative | negative | negative | negative | negative | negative | negative | negative | negative |

| PID | Ultra (sputum) semi-quantitative result | Ultra (swab 5) | Quantstudio (qPCR) results (storage temperature, storage duration) |  |  |  |  |  |  |  |  |  |  |  |
| --- | --- | --- | --- | --- | --- | --- | --- | --- | --- | --- | --- | --- | --- | --- |
|  |  |  | Swab 1 (2-8°C, 24 hours) |  |  | Swab 3 (2-8°C, 24 hours) |  |  | Swab 2 (-80°C, 1-5 months) |  |  | Swab 4 (-80°C, 1-5 months) |  |  |
|  |  |  | HL-1 | HL-2 | VBB | HL-1 | HL-2 | VBB | HL-1 | HL-2 | VBB | HL-1 | HL-2 | VBB |
| 25 | low | not detected | negative | negative | positive | negative | positive | negative | negative | negative | positive | negative | negative | negative |
| 26 | low | not detected | negative | negative | negative | negative | negative | negative | negative | negative | negative | negative | negative | negative |
| 27 | low | trace | positive | positive | positive | positive | positive | positive | positive | positive | positive | positive | positive | positive |
| 28 | low | not detected | negative | negative | negative | negative | negative | negative | negative | negative | negative | negative | negative | negative |
| 29 | low | trace | negative | negative | negative | negative | negative | negative | negative | negative | negative | positive | negative | negative |
| 30 | low | not detected | positive | negative | negative | negative | negative | positive | negative | negative | negative | negative | negative | negative |
| 31 | low | not detected | negative | negative | negative | negative | negative | negative | negative | negative | negative | negative | negative | negative |
| 32 | vlow | vlow | negative | negative | negative | negative | negative | negative | negative | negative | negative | negative | negative | negative |
| 33 | vlow | not detected | negative | negative | negative | negative | negative | negative | negative | negative | negative | negative | negative | negative |
| 34 | vlow | error 5007 | negative | negative | positive | negative | negative | negative | negative | negative | negative | negative | positive | negative |
| 35 | trace | not detected | negative | positive | positive | negative | negative | positive | negative | positive | positive | negative | negative | negative |
| 36 | trace | not detected | negative | negative | negative | negative | negative | negative | negative | negative | negative | negative | negative | negative |
| 37 | trace | not detected | negative | negative | negative | negative | negative | positive | negative | negative | negative | negative | negative | negative |
| 38 | trace | not detected | negative | negative | negative | negative | negative | negative | negative | negative | negative | negative | negative | negative |
| Total n/N (%) |  | 17/38 (45) | 13/38 (34) | 13/38 (34) | 18/38 (47) | 13/38 (34) | 13/38 (34) | 18/38 (47) | 15/38 (40) | 15/38 (40) | 19/38 (50) | 15/38 (40) | 10/38 (26) | 16/38 (42) |

HL-1, initial heat lysis; HL-2, repeat heat lysis; VBB, vortex bead beating; vlow, very low; quantitative polymerase chain reaction; error 5007 indicates insufficient sample processing or a failure in the assay's internal quality controls, positive indicates a MTBC-detected result by qPCR, negative indicates a MTBC not detected result by qPCR; invalid indicates that the assay control failed to amplify, Ultra refers to the Xpert MTB/RIF Ultra assay

### Phase Two

Among participants with valid results for all tests, all seven TS tested positive by qPCR in 3/39 (8%) participants (Table 2). When stored dry at −80°C, MTBC was detected in 9/20 (45%) specimens after 3-day storage compared to 11/20 (55%) after 7-day storage. When stored at −80°C in TE buffer, MTBC was detected in 8/19 (42%) and 10/19 (53%) TS specimens after 3-day and 7-day storage, respectively. When TS were stored dry at 37°C for 3 days, MTBC was detected in 10/19 (53%) specimens, compared to 7/19 (37%) after 7-day storage. MTBC was detected in 13/20 (65%) and 11/20 (55%) specimens, when stored in TE buffer at 37°C for 3- and 7-days, respectively. A comparison of Ct values for TS stored either “dry” or in TE buffer for 3 days, showed lower Ct values but greater variability when stored in TE buffer (Fig 4). When stored frozen for 7 days, TS stored “dry” or in TE buffer showed a similar range of Ct values while those stored at 37°C in TE buffer, showed lower Ct values with greater variability. Raw data can be found in supplementary materials (Table S2).

**Fig 4:**
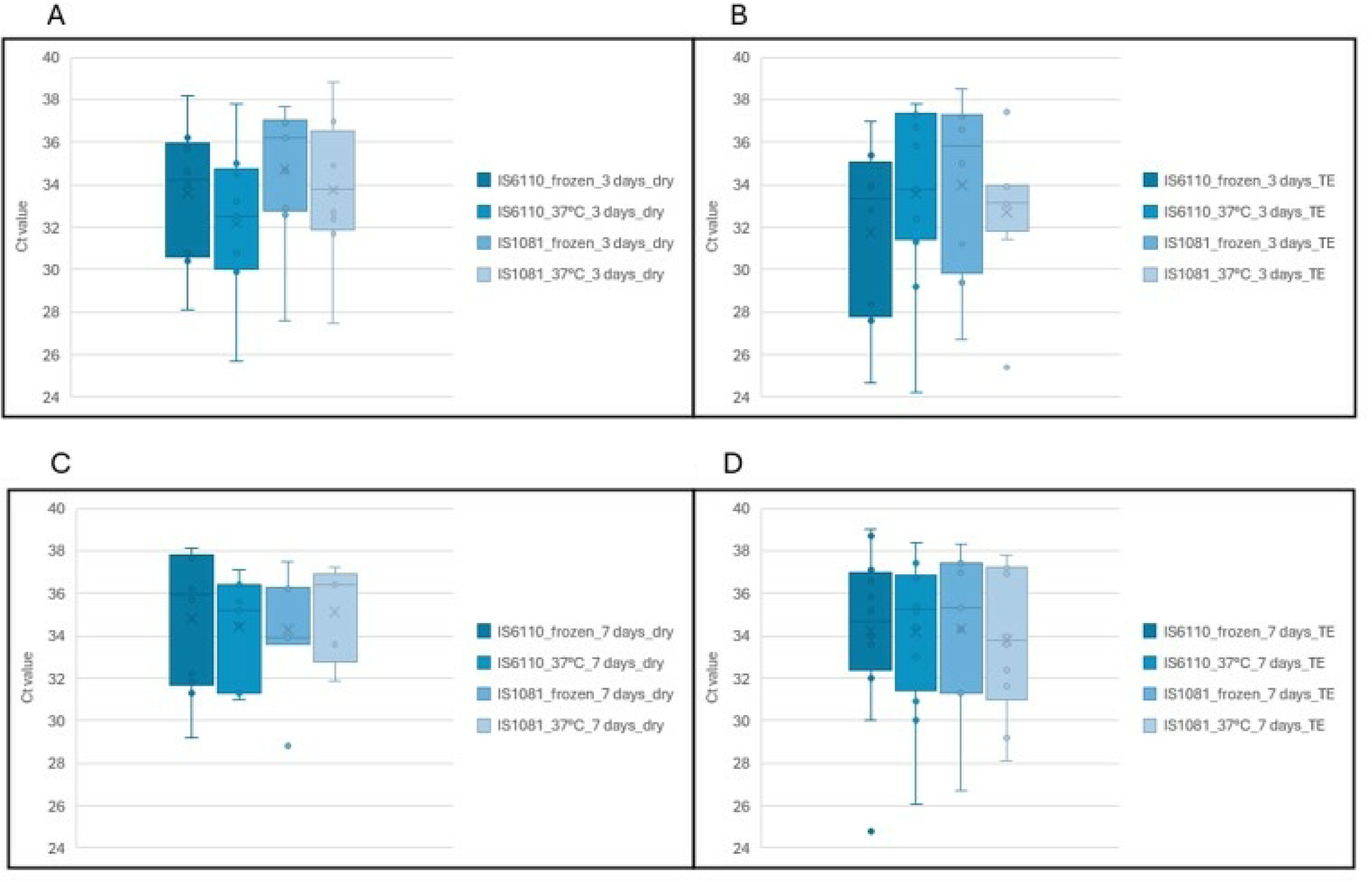
Comparison of Ct values for insertion elements (*IS*6110 and *IS*1081) obtained on qPCR between tongue swab specimens stored at different temperatures and durations. Figure 4A represents TS stored “dry” either frozen or at 37°C for 3 days; Figure 4B represents TS stored in TE buffer either frozen or at 37°C for 3 days; Figure 4C represents TS stored “dry” either frozen or at 37°C for 7 days and Figure 4D represents TS stored in TE buffer either frozen or at 37°C for 7 days

**Table 2:** Ultra semi-quantitative and qPCR results for tongue swabs from participants recruited during phase two.

| Participant number | Ultra semiquantitative result (sputum) | Ultra semiquantitative result (TS) PSM only | Ultra semiquantitative result (TS) PSM and SR | Quantstudio (qPCR) results (Storage temperature, transport condition, storage duration) |  |  |  |  |  |  |  |  |
| --- | --- | --- | --- | --- | --- | --- | --- | --- | --- | --- | --- | --- |
|  |  |  |  | 2-8°C, dry, 24 hours | -80°C, dry, 3 days | -80°C, dry, 7 days | -80°C, TE buffer, 3 days | -80°C, TE buffer, 7 days | 37°C, dry, 3 days | 37°C, dry, 7 days | 37°C, TE buffer, 3 days | 37°C, TE buffer, 7 days |
| 1 | high | medium | low | positive | positive | positive |  |  |  |  | positive | positive |
| 2 | high | trace | vlow | positive | positive | positive |  |  |  |  | positive | positive |
| 3 | high | error 2008 | low | positive | negative | positive |  |  |  |  | positive | positive |
| 4 | high | low | low | negative | positive | positive |  |  |  |  | positive | positive |
| 5 | high | low | low | positive |  |  | positive | positive | positive | positive |  |  |
| 6 | high | low | low | positive |  |  | positive | positive | positive | invalid |  |  |
| 7 | high | trace | not detected | negative | positive | negative |  |  |  |  | positive | positive |
| 8 | high | low | vlow | negative | positive | positive |  |  |  |  | positive | positive |
| 9 | high | not detected | trace | negative |  |  | positive | positive | positive | positive |  |  |
| 10 | high | low | vlow | negative | negative | positive |  |  |  | negative | positive | positive |
| 11 | high | low | trace | negative |  |  | positive | positive | positive | positive |  |  |
| 12 | high | low | low | negative |  |  | positive | positive | positive | positive |  |  |
| 13 | high | error 2008 | medium | positive | positive | positive |  |  |  | negative | positive | invalid |
| 14 | high | high | low | negative |  |  | positive | positive | positive | invalid |  |  |
| 15 | high | trace | not detected | negative |  |  | negative | negative | positive | positive |  |  |
| 16 | medium | vlow | trace | positive |  |  | positive | positive | positive | positive |  |  |
| 17 | medium | error 2008 | low | positive | positive | positive |  |  |  | negative | positive | positive |
| 18 | medium | vlow | trace | positive | positive | positive |  |  |  |  | positive | positive |
| 19 | medium | vlow | vlow | negative |  |  | positive | negative | positive | positive |  |  |
| 20 | medium | vlow | vlow | positive |  |  | negative | positive | negative | negative |  |  |
| 21 | medium | not detected | not detected | negative | negative | negative |  |  |  |  | positive | negative |
| 22 | medium | vlow | not detected | positive |  |  | negative | negative | negative | negative |  |  |
| 23 | medium | not detected | not detected | positive | negative | negative |  |  |  |  | negative | negative |
| 24 | medium | not detected | not detected | negative |  |  | negative | negative | negative | negative |  |  |
| 25 | low | not detected | not detected | negative |  |  | negative | negative | negative | negative |  |  |
| 26 | low | not detected | not detected | positive | negative | negative |  |  |  |  | negative | negative |
| 27 | low | vlow | not detected | negative |  |  | negative | positive | positive | negative |  |  |
| 28 | low | not detected | not detected | positive |  |  | negative | negative | negative | negative |  |  |
| 29 | low | error 2008 | not detected | positive |  |  | negative | negative | negative | negative |  |  |
| 30 | low | error 2008 | not detected | positive |  |  | negative | negative | negative | negative |  |  |
| 31 | low | not detected | not detected | positive |  |  | negative | negative | negative | negative |  |  |
| 32 | low | not detected | not detected | negative | negative | negative |  |  |  |  |  | negative |
| 33 | low | not detected | not detected | negative | positive | positive |  |  |  |  | negative | negative |
| 34 | vlow | not detected | not detected | negative |  |  | negative | positive | negative | negative |  |  |
| 35 | vlow | not detected | not detected | positive | positive | positive |  |  |  |  | positive | positive |
| 36 | trace | not detected | not detected | negative | negative | negative |  |  |  |  | negative | negative |
| 37 | trace | not detected | trace | negative | negative | negative |  |  |  |  | negative | negative |
| 38 | trace | not detected | not detected | negative | negative | negative |  |  |  |  | negative | negative |
| 39 | trace | not detected | trace | negative | negative | negative |  |  |  |  | positive | positive |
| **Total n/N (%)** |  | **18/39 (46%)** | **20/39 (51%)** | **18/39 (46%)** | **10/20 (50%)** | **11/20 (55%)** | **8/19 (42%)** | **10/19 (53%)** | **10/19 (53%)** | **7/19 (37%)** | **13/20 (65%)** | **11/20 (55%)** |

HL, heat lysis; VBB, vortex bead beating; vlow, very low; qPCR, quantitative polymerase chain reaction; MTM, molecular transport medium; error 2008 indicates a pressure related issue, positive indicates a MTBC-positive result by qPCR, negative indicates a MTBC not detected result by qPCR; Ultra refers to the Xpert MTB/RIF Ultra assay; invalid indicates that the assay control failed to amplify, cells without data indicate that testing was not performed under those storage conditions

### Ultra results

Swabs processed using MTM alone demonstrated MTBC detection on 18/39 (46%) participants with 5/39 (13%) specimens producing errors. When using a combination of MTM and SR buffer, MTBC was detected in 20/39 (51%) participants without any errors.

## Discussion

In this study, we assessed whether transport and storage conditions affect the performance of TS for detecting MTBC. In addition, we also modified processing methods in the laboratory to determine if assay sensitivity could be improved.

Phase 1 of the study demonstrated that MTBC can be detected from five serially collected swabs with 1-2 Ct value difference between TS from the same participant using HL and qPCR. However, results also demonstrated that MTBC detection can be sporadic, especially on those participants with a lower bacillary load, as determined by the Ultra sputum semi-quantitative result. The addition of VBB improved detection of MTBC and produced lower Ct values compared to HL alone. Storage of TS at −80°C for up to 5 months, showed similar detection of MTBC, compared to testing within 24 hours after 2-8°C storage. Phase 1 also demonstrated that TS were compatible with Ultra, but we believed that assay sensitivity could be improved using protocol modification.

For phase 2, a combination of HL and VBB was used for TS pre-processing before testing on qPCR. Although specimen numbers were small and storage duration was limited (up to 7 days), when stored at −80°C, MTBC was detected in a similar number of participants when stored “dry” or in TE buffer. When stored “dry,” for 3-days, there was improved MTBC detection and lower Ct values when TS were stored at 37°C compared to TS stored at −80°C. When stored in TE buffer for 3 days, there was improved MTBC detection on TS stored at 37°C but lower Ct values observed on TS stored at −80°C. When stored for 7-days, “dry” storage at −80°C demonstrated improved detection of MTBC with similar Ct values obtained compared to TS stored at 37°C. For TS stored in TE buffer for 7 days, there was similar MTBC detection and similar Ct values observed between TS stored at 37°C and −80°C. A storage duration beyond seven days was not evaluated, as this typically exceeds the time specimens take to reach laboratories in South Africa (7). Additionally, prolonged storage, at room temperature or higher, albeit for respiratory specimens are prone to overgrowth by contaminants, compromising downstream testing (8).

When using SR buffer alone, the TS Ultra error rate was high indicating a specimen processing issue, however, MTBC was detected in 45% of participants, showing TS combability with the assay. To enhance detection, Phase 2 evaluated two additional methods: one using MTM alone and the other combining MTM with SR buffer. MTM has been shown to improve MTBC detection in sputum with low bacillary loads while also stabilizing specimens for transport (9). However, it was hypothesized that SR buffer might be necessary for optimal assay performance. Given that TS do not require the same liquefaction treatment as sputum, concerns arose that undiluted SR buffer could be too harsh. Therefore, a diluted SR buffer was tested to assess its impact on TS processing. MTBC was detected in 46% of TS transported in MTM only with an error rate of 13% while detection improved to 51%, with no errors produced, when a combination of MTM and SR buffer was used suggesting that our hypothesis was correct in that SR buffer may be required to for processing of TS prior to testing on Ultra. Although the buffer combination with MTM improved detection, it was subsequently found to be difficult to procure MTM in SA. There was also the consideration of the increased cost associated with purchasing MTM compared to transporting a TS without any buffer.

While this was not a comprehensive study and definitive conclusions cannot be drawn, study findings suggest that TS can be transported in TE buffer at temperatures up to 37°C for ∼3 days without compromising sample integrity. For long-term preservation, storage at −80°C (either “dry” or in TE buffer) appears to offer greater stability. Additionally, TS may be transported and stored “dry”, depending on logistical and programmatic considerations.

This research provided essential insights that informed the design of larger TS clinical performance evaluation studies performed by our group, helping to optimize TS testing for MTBC detection.

## Conclusions

This study provides evidence that TS can be used for MTBC detection, with diagnostic yield influenced by transport, storage, and processing conditions. Key optimizations, such as vortex bead beating, diluted SR buffer, and flexibility in short-term storage at 37°C or long-term storage at −80°C, enhanced detection while maintaining specimen stability. Although MTM improved performance, its limited availability and higher cost may pose barriers in resource-constrained settings. These findings offer practical insights to inform future implementation and scale-up of non-sputum-based TB diagnostics, supporting efforts to reach populations for whom sputum collection is difficult or inadequate.

## Data Availability

All relevant data are within the manuscript and its Supporting Information files.

## Acknowledgements

The authors would like to thank Vidya Keshav for assistance with editing, the study participants and the Johannesburg Health District and the and the Gauteng Department of Health for their support and collaboration on this study.

## Supporting information

Table S1: Raw qPCR results and Xpert MTB/RIF Ultra results from phase one

Table S2: Raw qPCR results and Xpert MTB/RIF Ultra results from phase two

